# How Good Are Large Language Models at Supporting Frontline Healthcare Workers in Low-Resource Settings – A Benchmarking Study & Dataset

**DOI:** 10.1101/2025.08.26.25333975

**Authors:** Samuel Rutunda, Gwydion Williams, Kleber Kabanda, Francis Nkurunziz, Solange Uwiduhaye, Eulade Rugegamanzi, Cyprien Nshimiyimana, Vaishnavi Menon, Mira Emmanuel-Fabula, Alastair K. Denniston, Xiaoxuan Liu, Emery Hezagira, Bilal A. Mateen

**Author notes:** Joint first authors (Samuel Rutunda, Gwydion Williams). <u>**Corresponding Author:**</u> Prof. Bilal A Mateen, 437 N 34th Street, Seattle, WA 98103, USA.

## Abstract

Large language models (LLMs) have demonstrated strong performance in medical contexts; however, existing benchmarks often fail to reflect the real-world complexity of low-resource health systems accurately. This study developed a dataset of 5,609 clinical questions contributed by 101 community health workers (CHWs) across four Rwandan districts and compared responses generated by five large language models (LLMs) (Gemini-2, GPT-4o, o3 mini, Deepseek R1, and Meditron-70B) with those from local clinicians. A subset of 524 question-answer pairs was evaluated using a rubric of 11 expert-rated metrics, scored on a five-point Likert scale. Gemini-2 and GPT-4o were the best performers (achieving mean scores of 4.49 and 4.48 out of 5, respectively, across all 11 metrics). All LLMs significantly outperformed local clinicians (ps < 0.001) across all metrics, with Gemini-2, for example, surpassing local GPs by an average of 0.83 points on every metric (range: 0.38 – 1.10). While performance degraded slightly when LLMs communicated in Kinyarwanda, the LLMs remained superior to clinicians and were over 500 times cheaper per response. These findings support the potential of LLMs to strengthen frontline care quality in low-resource, multilingual health systems.

---

Large language models (LLMs) have consistently demonstrated expert-level performance on postgraduate medical examinations such as the USMLE^1^, and in navigating clinical vignettes that approximate real-world scenarios with similar levels of accuracy^2^. However, these assessments fail to reflect the complexities of tiered health systems commonly found in low- and middle-income countries (LMICs). In these settings, frontline care is often delivered by narrowly trained community health workers (CHWs); diagnostic and therapeutic resources may be scarce; and healthcare delivery frequently occurs in non-English language environments, posing additional linguistic challenges^3^.

The development of the AfriMedQA dataset addressed a critical gap by creating the world’s first large-scale English-language African medical multiple-choice question (MCQ) dataset^4^. The accompanying benchmarking study revealed performance differences on ‘African’ questions compared to MedQA/USMLE-derived questions^5^. The significance of such datasets lies not only in representational equity but also in their ability to encode the meaningful differences in disease burden, clinical presentation, and healthcare infrastructure that likely explain (in part) the observed geography-based variation in LLM performance. However, benchmarking datasets that mirror the realities of healthcare in resource-limited settings remain scarce. This scarcity hinders our understanding of whether existing LLMs are suitable for these contexts and limits opportunities to de-risk deployments through in-silico testing.

To address this, we engaged 101 CHWs across four Rwandan districts (Gicumbi, Gakenke, Nyanza, and Ngoma) to generate open-ended clinical questions (i.e, vignettes) based on typical patient encounters. CHW demographic characteristics are detailed in Supplementary Table 1. Participants were encouraged to submit at least 60 questions over three weeks via a custom data collection app (‘Mbaza’), developed by Digital Umuganda, a Rwandan technology company. Questions were submitted as voice recordings in Kinyarwanda, transcribed using a speech-to-text model developed by Digital Umuganda^6^, and subsequently cleaned and screened for quality and relevance by trained local nurses.

Out of 7,143 questions submitted, 1534 were excluded based on quality criteria, resulting in 5,609 accepted entries. Questions were mapped to 18 domains that aligned with the 14 government-defined CHW work packages (Supplementary Figure 3). Multiple category assignments were allowed per question. The most common category was “Other” (n = 1,613), followed by “Malaria” (n = 1,133) and “Maternal and Newborn Health” (n = 802). The least frequent categories included “Emergency Response to Epidemics” (n = 56), “Gender-Based Violence” (GBV) (n = 72), and “Adolescent and Sexual Reproductive Health” (n = 215). Quality assessment and categorisation were completed by a group of local nurses (demographics in Supplementary Table 2).

Following data collection, a group of Rwandan general practitioners and senior nurses (demographics provided in Supplementary Table 3) was recruited to generate clinician responses to each question, simulating the ideal response a Community Health Worker (CHW) might receive if such a ‘tele-advice’ service existed. Questions and clinician responses were translated into English or Kinyarwanda to create a fully bilingual dataset. These (translation) outputs were subsequently reviewed and edited by professional linguists. In parallel, responses to the questions were generated using five LLMs: Gemini-2-Flash, GPT-4o, o3-mini-high, Deepseek-R1, and Meditron-70B. An illustrative example of a question, along with a paired clinician and LLM response, is provided in Box 1.

**An illustrative (mental health-related) query and a pair of responses (clinician & LLM). Note that the individuals depicted and discussed here are illustrative, and are not real people.**

**Question from CHW**

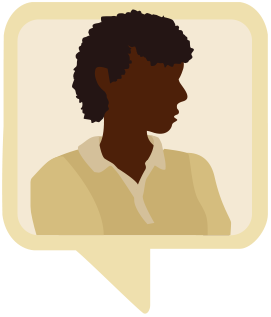

“A fifty-two-year-old woman, weighing thirty-eight kilograms, has been speaking loudly during our discussions to commemorate the genocide against the Tutsi in 1994. I have taken her to a quiet place. Help me. Since I see she has a trauma issue, what other help can I provide her?”

**Response from Clinician**

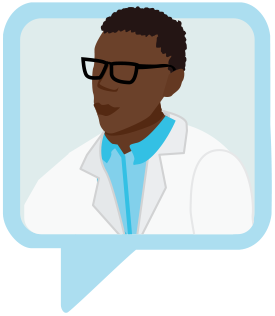

“Well, if you have really failed to make her come back to normal state, then that woman might be having a psychiatric condition (mental disorder) called Post traumatic stress disorder(PTSD) that happens to people who have encountered multiple traumatic events that occurred anytime in the past. and also most patients with PTSD have other mental disorder like Major depressive disorder, anxiety disorder, bipolar disorder, therefore she needs a psychiatrist for better management that includes pharmacotherapy & psychotherapy.”

**Responsefrom Gemini-2 (LLM)**

“Okay, Murakoze cyane (thank you very much) for recognizing her distress and taking her to a quiet place. That was a very important first step. It’s understandable that the commemorations can trigger traumatic memories, especially considering the weight of the past. Given her age and low weight, and the context of the commemorations, let’s consider a few things:

**IMMEDIATE ACTIONS**

**Safety and Comfort:** Make sure she feels safe and secure. Your presence and calm demeanor are already helpful. Offer her water and a comfortable place to sit or lie down.

**Grounding Techniques:** Encourage her to focus on her senses.

You can ask her:

“What are five things you can see?”

“What are four things you can touch?”

“What are three things you can hear?”

“What are two things you can smell?”

“What is one thing you can taste?”

**Breathing Exercises:** Guide her in slow, deep breathing. Encourage her to breathe in deeply through her nose, hold it for a few seconds, and exhale slowly through her mouth. This can help calm her nervous system.

**FURTHER SUPPORT AND REFERRAL:**

**Mental Health Services:** This is crucial. Rwanda has made efforts to provide mental health services. Find out what resources are available in your community. This might include:

1. **Health Centers (Ibitaro Nto):** Many health centers have trained nurses who can provide basic mental health support.
2. **District Hospitals (Ibitaro by’Akarere):** These hospitals typically have doctors and mental health professionals.
3. **Specialized Mental Health Facilities:** If possible, referral to a specialized facility might be necessary. Inquire about the closest options.
4. **Community-Based Programs:** Are there any local organizations offering trauma counseling or support groups?

**Nutritional Support:** Given her low weight, assess her nutritional status. Is she able to access food regularly? Connect her with resources like food distribution programs or agricultural support initiatives, if available.

**Social Support:** Connect her with her family, friends, or community members who can offer ongoing support. Social isolation can worsen trauma symptoms.

**Explain Confidentiality:** Assure her that what she shares with you will be kept confidential, unless there is a risk of harm to herself or others.

**ADDRESSING POTENTIAL TRAUMA:**

**Listen Without Judgment:** Allow her to share what she’s comfortable sharing, without pressure. Do not interrupt or offer unsolicited advice. Just listen attentively and acknowledge her feelings. A simple “Ndashima ko ubimbwira (Thank you for telling me)” can be powerful.

**Avoid Triggering Questions:** Be very careful not to ask specific questions about the genocide events. Let her lead the conversation.

**Validate Her Feelings:** Tell her it’s okay to feel the way she’s feeling. Her reactions are understandable given the circumstances and her past experiences. For example, you could say, “Birumvikana ko wumva utyo (It’s understandable that you feel that way).”

**Check for Physical Needs:** Her loud speaking and distress could also be related to physical needs. Is she hungry, thirsty, or in pain? Given her low weight, she might be malnourished, which can exacerbate emotional distress.

**IMPORTANT CONSIDERATIONS FOR RWANDA:**

**“Kwibuka” (Remembrance):** Be aware of the specific cultural sensitivities surrounding the genocide commemorations.

**“Gacaca” Courts Legacy:** The Gacaca courts, while aiming for reconciliation, may have also caused further trauma for some. Be mindful of this history.

**Traditional Healing:** While referring to formal mental health services, also be respectful of traditional healing practices that may be helpful to her.

**Resource limitations:** Given her low weight, check if she has access to food. If not refer her to a nutritionist and find out whether there are any options for helping her access basic food.

**Safety and fairness:** Be careful when talking to the patient to observe confidentiality. Do not do anything to aggravate her situation.

**SELF-CARE FOR YOU:**

Supporting someone experiencing trauma can be emotionally draining. Remember to take care of yourself. Talk to a colleague or supervisor about your experience and seek support if needed.

What specific resources do you have access to in your community? Knowing that will help me give you more targeted advice. This is a difficult situation, and you are doing a great job by being present and supportive. Keep up the compassionate work!”

A random subset of 524 question-response pairs was selected for expert evaluation (demographics for expert evaluators in Supplementary Table 4). Evaluators assessed each response (generated by clinicians and the five LLMs) using an 11-item rubric based on the framework introduced in Google’s Med-PaLM-2 evaluation^1^, employing a five-point Likert scale (see Online Methods). The first 416 questions were evaluated based on the prompts and responses in English. The remaining 108 were conducted entirely in Kinyarwanda. Any unresolved disagreement between evaluators (defined as a difference >1 between scores given by evaluators for the same question-response pair) would lead to affected questions being removed from the final dataset 15 English questions and 3 Kinyarwanda questions were removed from the final dataset on this basis, yielding a final set of 506 questions. The results of the evaluation are presented in Figures 1a and 1b.

**Figure 1.**
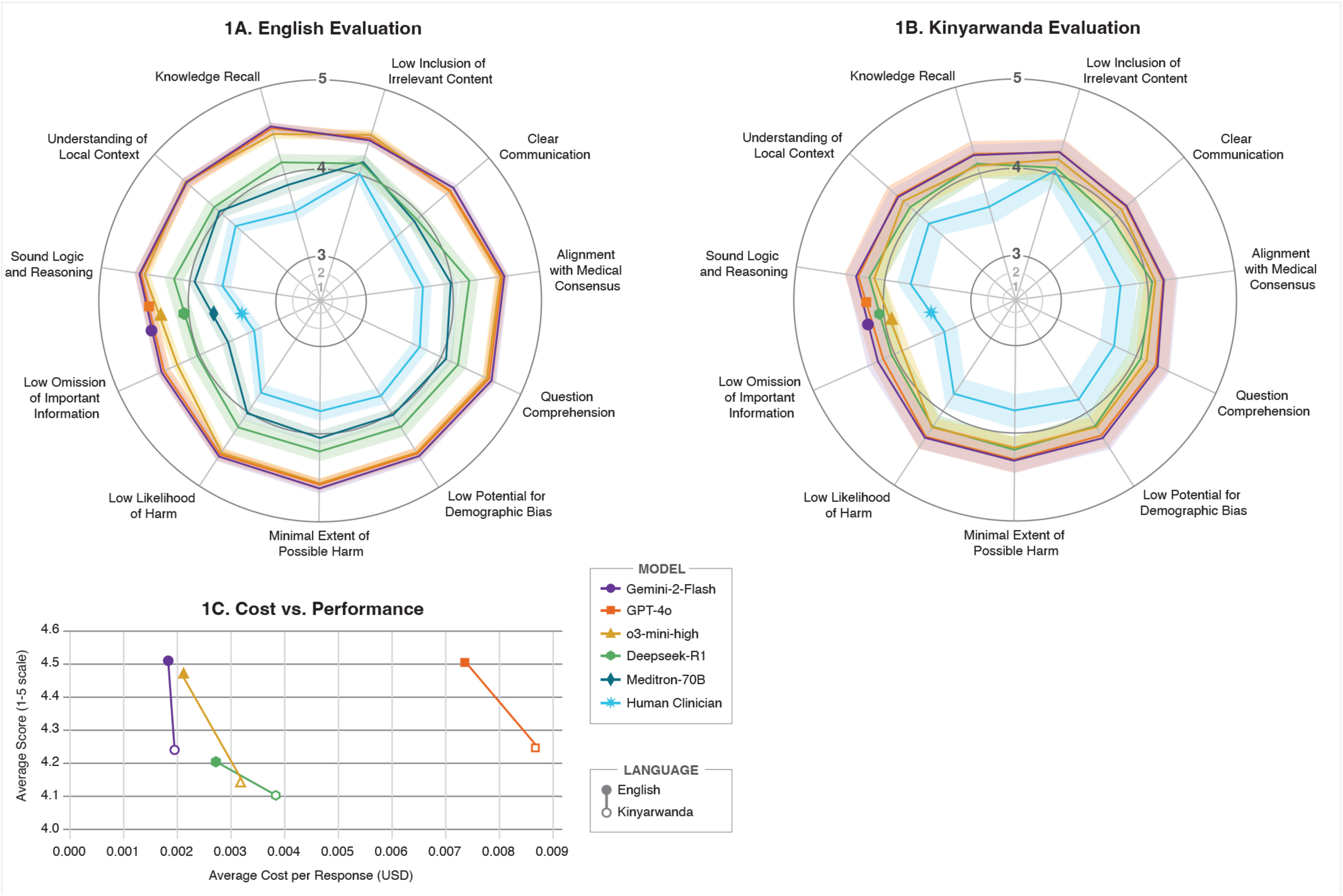
Evaluation scores (and costs) for all models and clinicians on the 11 evaluation criteria when prompted in English & Kinyarwanda. Figure la (top left) and 1b (top right), each bold line represents the mean score, with the shaded region of the same colour representing the 95% confidence interval. The maximum score is 5, and the minimum is 1. Figure 1c (bottom left) illustrates the (average) cost (per response) versus average score across all evaluation metrics for Gemini-2-Flash, o3-mini-high, Deepseek-RI. and GPT-4o. Meditron-70B is excluded from 1b since it cannot ‘ speak” Kinyarwanda and from the cost analysis since it does not have a standardised token cost given that it is open-source, and not accessed via API, i.e., the costs are almost entirely a function of the compute costs which can be profoundly different if a local/edge solution can be configured rather than utilising cloud compute. Human clinicians are excluded from 1c given their costs are several orders of magnitude greater than even the most expensive model.

When prompted in English, the top-performing LLMs, Gemini-2 and GPT-4o, achieved average performance scores (out of 5) of 4.56 (SD 0.58) and 4.53 (SD 0.68), respectively. The o3 mini model performed comparably on most metrics (mean 4.49, SD 0.58) but underperformed on ‘omission of important information’ (falling 0.21 and 0.19 points behind Gemini-2 and GPT-4o, respectively). Deepseek R1 (mean 4.16, SD 1.06) and Meditron-70B (mean 3.99, SD 0.86) had markedly lower performance. Pairwise comparisons between all models and human clinicians are summarised in Supplementary Figure 1. Notably, all LLMs significantly outperformed local clinicians (ps < 0.001) across all metrics, with Gemini-2, for example, surpassing local GPs by an average of 0.83 points (range: 0.38 – 1.10). Evaluators appeared to favour the LLMs’ structured and comprehensive responses over the clinicians’ briefer answers. This brevity likely contributed to clinicians scoring well on ‘absence of irrelevant content’ (GPs: mean 4.02, SD 0.99; nurses: mean 3.99, SD 0.99) but poorly on ‘omission of important information’ (GPs: mean 3.38, SD 0.91; nurses: 3.28, SD 0.89).

Performance varied by CHW work package. Clinicians demonstrated the most significant variation between their best (GPs: Water, Sanitation, and Hygiene: 3.99; nurses: Maternal & Newborn Health: 4.08) and worst (GPs & nurses: Family Planning, 3.42 & 3.24) topics. In contrast, Gemini-2 exhibited only a 0.31-point drop from its highest (Mental Health, 4.63) to its lowest (Water, Sanitation, and Hygiene, 4.32) scoring topics. Frequencies of each topic area within the 506-question subset are provided in Supplementary Figure 4; corresponding performance data are in Supplementary Figure 5.

For the 105 Kinyarwanda-prompted questions, Meditron produced unusable outputs; therefore, it was excluded from this analysis. Of the remaining four models, performance decreased by a mean of 0.15 points across all metrics compared to their English-prompted outputs (Figure 1), yet remained superior to clinician responses (ps < 0.001; Supplementary Figure 2). Performance data per LLM, metric, and language (including all relevant pairwise comparisons) are provided in Supplementary Figures 1, 2, and 7. Topic-level performance trends were consistent with the English subset.

Clinicians had the option to respond in either English or Kinyarwanda, based on personal preference. Language preference broadly aligned with professional background; general practitioners preferred English (likely due to their language of training), while nurses favoured Kinyarwanda (see Supplementary Table 3). We observed no significant differences in the scores received by clinicians who opted to respond in English versus Kinyarwanda (p = 1.000).

Overall, the study demonstrates that LLMs can provide high-quality, on-demand clinical advice to CHWs that outperforms local experts, even when operating in low-resource, non-English language settings. However, it is worth remembering that the workflow here (i.e., Q&A) does not fully reflect the complexity of day-to-day practice, nor does it guarantee that other human factors (e.g., CHWs not complying with the advice) would not undermine the translation of these results into patient-level benefits if an LLM-based clinical decision support system were to be deployed in this manner.

The latter result, pertaining to the language in which the LLM was prompted, aligns with prior findings showing performance improvements when prompting LLMs with high-quality English translations instead of less-well-represented languages^7^. However, the additional cost and latency associated with integrating professional linguists or machine translation APIs must be considered. For many use cases, the modest performance reduction related to native language input may be offset by the operational advantages of direct Kinyarwanda interaction.

A cost analysis highlights the economic benefits of LLMs. Clinician-generated answers cost an average of $5.43 (general practitioners) or $3.80 (nurses) per question – likely an overestimate due to consulting premiums. In contrast, LLM responses cost an average of $0.0035 in English and $0.0044 in Kinyarwanda, which we found to be a significant increase (p <.001; illustrated in Figure 1c). This was driven by significantly higher (p <.001) token counts per response in Kinyarwanda (mean of 1173) than in English (mean of 905), despite similar word counts and semantically identical queries. This is consistent with prior findings that non-English languages use more tokens for equivalent content^8^, and highlights the need for improved tokenisation methods for less well-represented languages.

Finally, while human expert evaluations remain the gold standard for research-based LLM assessment in healthcare, our findings highlight their long-term unsustainability as an oversight mechanism for real-world deployments. Even if we use 10% of the evaluator’s costs (who were paid $9.17 per assessment), these costs become impractical at scale. Considering a national-level rollout, for example, Rwanda has ~60,000 CHWs; even one question every working day from every CHW would cost over $13 million per annum to quality assure. This doesn’t even factor in the potential impact of multi-turn interactions. Future research should prioritize the development and validation of automated evaluation strategies^9^, and explore how these can be operationalized for real-time performance monitoring post-deployment.

In conclusion, these results suggest great promise for LLM-based clinical decision support tools in supporting frontline healthcare workers to deliver a higher standard of care in low-resource settings. Confirmation of this potential role for LLMs requires in-field studies, and prospective evaluation of the impacts on healthcare outcomes, which are currently underway^10,11^.

## Online Methods

This study was conducted in four districts across Rwanda: Gicumbi (Byumba Health Centre) and Gakenke (Nganzo Health Centre) in the Northern Province, Nyanza (Nyanza Health Centre) in the Southern Province, and Ngoma (Kibungo Health Centre) in the Eastern Province. Below, we outline the process by which we generated the benchmarking dataset and describe our evaluation of both human and model performance.

### Dataset Generation

Dataset generation happened in four phases. First, we recruited Rwandan community health workers (CHWs) to generate vignettes that captured representative cases they would encounter in the field. Second, local nurses assessed the quality of those vignettes (and rejected any that failed to meet set standards) and categorised them by health area. Third, local linguists translated approved vignettes from Kinyarwanda into English. Fourth, and finally, local clinicians generated responses to the vignettes, which were themselves translated to provide bilingual (English & Kinyarwanda) responses. Below is a more detailed explanation of each step.

#### 1. Vignette Generation by CHWs

To generate vignettes representative of patients encountered by frontline health workers in Rwanda, we recruited 101 community health workers (CHWs) across four districts (demographic data for CHWs by county are provided in Supplementary Table 1). CHWs were contacted and recruited based on recommendations from the Rwanda Biomedical Centre (RBC), the para-statal implementation arm of the Ministry of Health, which manages the community health programme. Specifically, we relied on input from the head of the community health programme at RBC, Dr Emery Hezagira (also an author of this manuscript). Participation was voluntary, and CHWs were not paid directly for their time. However, all travel costs to and from training (see below) were covered, and smartphones were provided to all participating CHWs to support data collection.

Once recruited, all participating Community Health Workers (CHWs) were invited to a district-specific training workshop held in December 2024. During these one-day workshops, participants were trained to generate vignettes using an adaptation of the ‘Situation, Background, Assessment, and Recommendation’ (SBAR^12^) framework. This involved instructing CHWs to describe how a patient presented (situation, e.g., their symptoms, age, gender, and weight), any relevant contextual information or clinical history (background, e.g., any relevant pre-existing conditions), their analysis of the situation and the options they considered in response (assessment), the actions they took or recommended others take (recommendation), and any questions they have regarding the case for trained clinicians. CHWs were instructed to submit their vignettes via a custom-built mobile application, ‘Mbaza’, developed by Digital Umuganda specifically for this project. Vignettes were to be submitted via voice recording, and to ensure that recorded vignettes were of a high quality, CHWs were given the following additional instructions:

1. Background noise check: Ensure that you are in a quiet environment with no background noise in the audio.
2. Microphone check: Ensure that your microphone is properly working.
3. Medical Categorization: Ensure that all questions asked are within the 14 work packages/18 clinical domains.
4. Language & Terminology Standardization: Ensure that the terminology used in the questions is clear, concise, and consistent with local healthcare practices and languages.
5. Clarity: Make sure that the questions are clear and understandable by both the LLM and the General practitioners/nurses who will respond.
6. Completeness: Ensure the questions are complete and do not lack essential details such as the patient’s age, gender, and name of the disease/issue.

All training (including the above instructions) were delivered in Kinyarwanda, and the content provided here has been translated into English for documentation purposes.

Following training, all participating Community Health Workers (CHWs) generated vignettes by submitting voice recordings while working in their communities over a three-week period, with each CHW aiming to produce at least 60 vignettes (though many exceeded this target). This yielded 7143 vignettes to be assessed and categorised in the next stage (described below).

#### 2. Assessment & Categorisation by Nurses

Six nurses (three male, three female) were recruited to assess and categorise the 7,143 vignettes generated by all participating CHWs. To be eligible to participate, nurses needed (i) to have at least 3 years of clinical experience in community health and patient management, (ii) to be bilingual English-Kinyarwanda speakers, and (iii) basic familiarity with digital devices and applications. Nurses were recruited by a senior doctor based at the Butaro District Hospital, who was previously known to Digital Umuganda and leveraged their network to advertise the opportunity (author SU). Interested nurses then completed an application form, and those who were deemed eligible were recruited to the study. All participating nurses were paid 697 Rwandan Francs (~0.48 USD) per vignette processed.

Once recruited, nurses received targeted training sessions (which were delivered by Digital Umuganda over 2 days) on how to assess and categorise vignettes, which included practical simulations to ensure uniformity in approach. For each vignette, nurses would listen to the original audio recording and review a machine-translated transcript of the recording (transcripts were generated using the Digital Umuganda-maintained Mbaza speech-to-text model, and nurses could correct transcriptions upon review) before assessing the vignette for quality. Nurses were instructed to reject vignettes from inclusion in the final dataset if they failed to follow the SBAR tool described above, lacked sufficient information for a clinician to provide a sound response to the question posed, or if the audio recording was incomplete or inaudible. Nurses were not asked to record the reason for exclusion. A total of 1534 vignettes were rejected, leaving 5609 for categorisation. Nurses would then categorise each vignette that passed quality assessment into one/more of 18 medical domains, which align with the 14 ‘work packages’ that Rwandan CHWs are expected to provide care (including an “*Other*” option was also available if the vignette included elements that did not fit neatly within the 17 explicit domains). All 5,609 vignettes were categorised, and the distribution of vignettes per category is shown in Supplementary Figure 3. This was all completed within a custom-built annotation platform (which again was developed by Digital Umuganda specifically for this project) and was completed over 3 weeks.

#### 3. Transcription & Translation by Linguists

To facilitate the accurate transcription of the 5,609 categorised vignettes, we recruited eight linguists who would work under the supervision of two supervising linguists with previous experience of working with Digital Umuganda on English-Kinyarwanda NLP workflows. The two supervising linguists were existing collaborators of Digital Umuganda, and they shared the opportunity to participate in the study with other linguists in their network. Interested linguists completed an application form, which included assessments of their ability to conduct bidirectional English-Kinyarwanda translation. The eight highest-scoring applicants (as judged by the supervising linguists) were recruited for the study. Linguists were paid 1,629 Rwandan Francs (~$ 1.13 USD) per vignette reviewed.

Initial speech-to-text transcription was performed using Digital Umuganda’s Kinyarwanda speech-to-text model^6^. All transcriptions were then reviewed by the eight linguists, who listened to the audio recording while reviewing the transcribed text and correcting any errors that they found. Supervising linguists then reviewed a randomly sampled 10% of all transcripts reviewed by each linguist to ensure quality and consistency. This process was completed within a custom-built web-app developed by Digital Umuganda.

Verified transcriptions were initially translated using Digital Umuganda’s machine translation tool (Mbaza MT)^13^. The first 2784 questions were translated using Mbaza MT. It was then determined that GPT4o was capable of effectively translating the text; thus, the remainder of the questions were translated using this tool.

GPT-4o was prompted with the following text:

> *“Provide only the direct translation of the text below. Do not include any explanations, notes, or additional context*.*”*.

This prompt was included because although Gemini-2 anecdotally produced better translations, it was incapable (despite prompting) of not responding to the questions alongside the translation, which GPT4o (with prompting) was capable of doing.

Translations were verified by our team of linguists using the same procedure as for transcription verification: they compared the Kinyarwanda transcription with the English translation, listened to the original audio recording if necessary to resolve any misunderstandings, and corrected any errors. Supervising linguists again reviewed a randomly sampled 10% of the translations reviewed by each linguist to ensure quality and consistency. For the original Mbaza MT solution, out of the 2,784 translations examined, only 586 were not edited or corrected by the linguists at all. For GPT4o, 2711 were not modified by the linguists, suggesting a much higher quality initial output. Given that every translation was reviewed and, where necessary, edited by a linguist, we have a strong prior that the tool used should not impact the downstream assessment of the responses.

This process yielded linguist-verified transcriptions and translations of all 5,609 vignettes, providing a fully bilingual dataset of real questions generated by Rwandan community health workers (CHWs).

#### 4. Response Generation by Clinicians

Finally, to generate responses to the questions posed in the vignettes, we recruited six senior nurses with at least five years of clinical experience and 14 general practitioners (GPs) with 2 to 5 years of clinical experience. All nurses were recruited by the same senior doctor, who managed the recruitment process for vignette assessment and categorization, using the same procedure, but with more stringent eligibility criteria (i.e., requiring that participating nurses had at least 5 years of clinical experience). General practitioners (GPs) were recruited by the Director of Clinical Services at the Butaro District Hospital (author ER), with all participating GPs working at the same hospital. All senior nurses and GPs were paid 5,488 Rwandan Francs (approximately 3.80 USD) and 7,835 Rwandan Francs (approximately 5.43 USD), respectively, per vignette answered.

A dedicated web platform was created by Digital Umuganda to facilitate response generation, as for the other activities (see mFigure 1). Clinicians received each vignette and question in both audio and text formats, with the text format available in both Kinyarwanda and English. Upon accessing the question, clinicians first selected their preferred language for responding (responses could be given in either English or Kinyarwanda). Once selected, the audio recording in Kinyarwanda could be played, and the corresponding text was displayed in their chosen language. See Supplementary Table 3 for the distribution of preferred clinical language.

**mFigure 1.**
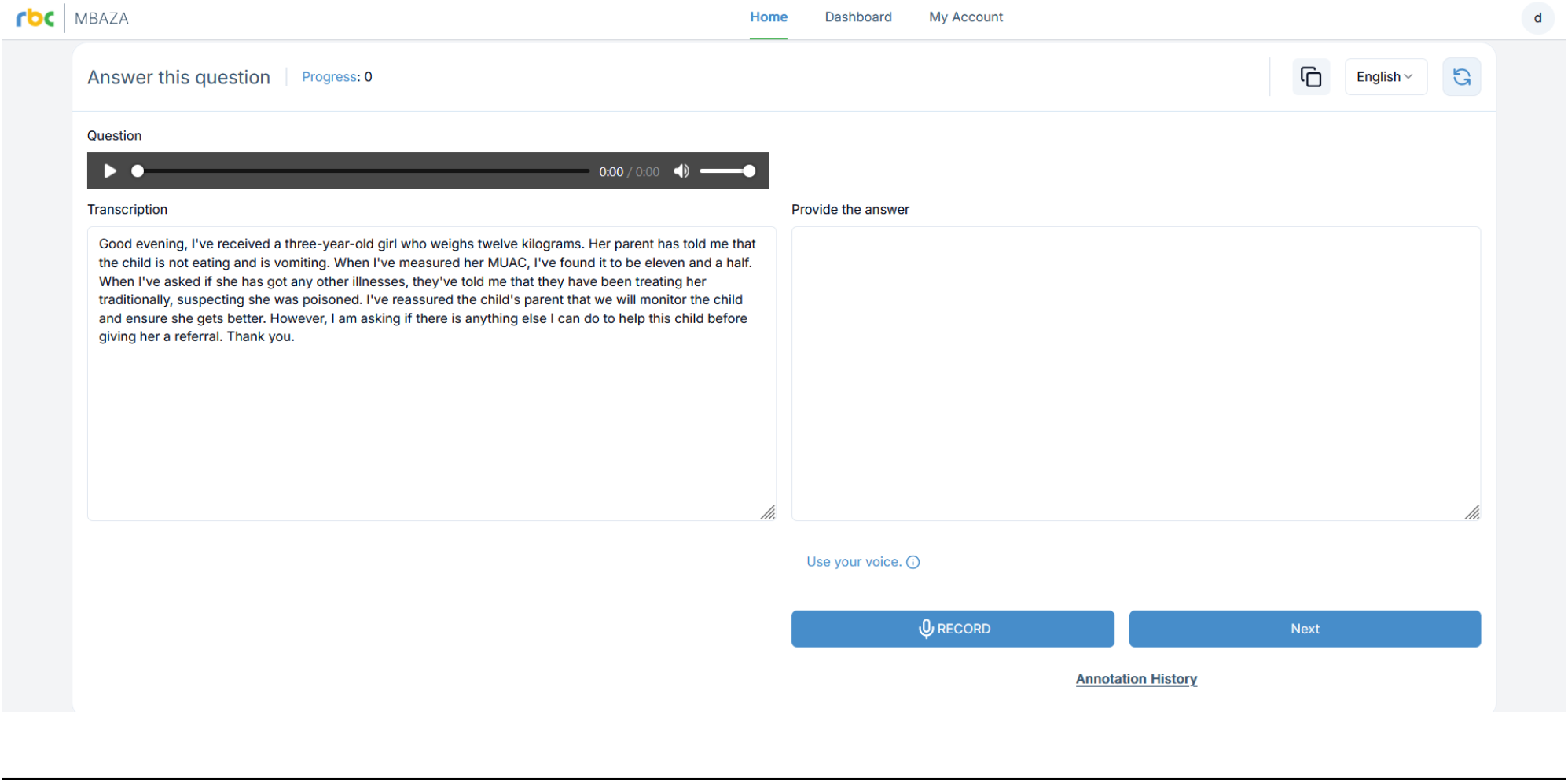
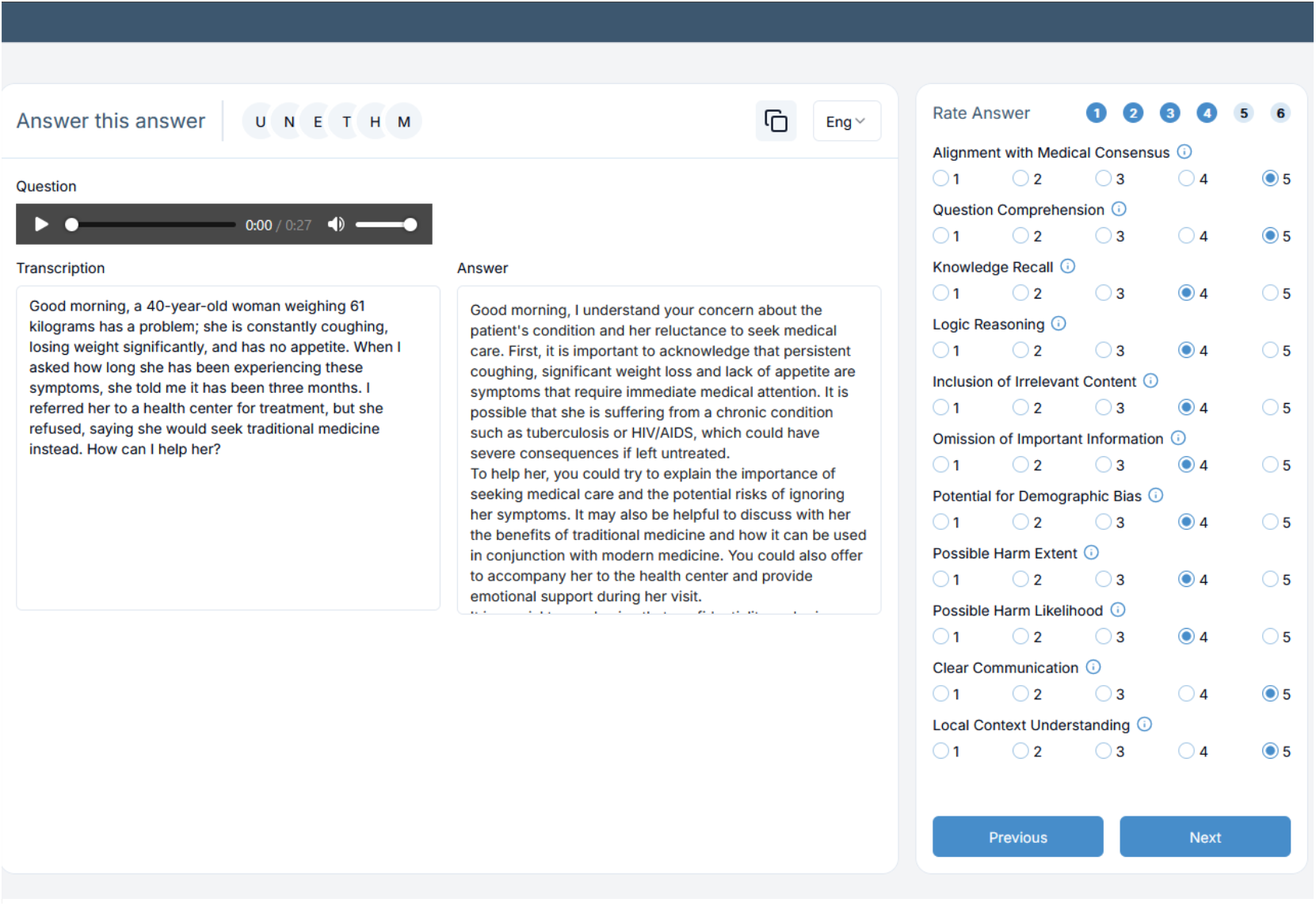
The *Response and Evaluation Web Application* Legend: The Response and Evaluation Web Portal presented clinicians with the original audio recording of the question alongside the transcription of that recording in a language of their choosing (Top). Having listened to and read the question, clinicians would respond by either typing in the text box provided or by recording their speech (which could later be edited). Once happy with their response, they would click Next, and then be prompted to rate the question along four dimensions (Relevance, Clarity, Actionability, and Completeness). The same platform was used for the human expert evaluation described later (Bottom). Note that the people discussed here are illustrative, and do not correspond to real cases/patients.

Clinicians were encouraged to listen to the original voice recording before responding to it. To ensure they understood the vignette, clinicians were asked to summarise the question posed in their own words before providing a detailed response. To facilitate ease of use, the platform included a speech-to-text model, allowing clinicians to dictate their responses, which would then be automatically transcribed (and clinicians could then edit). Finally, once submitted, each response was translated into the alternate language (again using GPT-4o for translation). This process yielded a fully bilingual set of 5,422 question-answer pairs – see mTable 1 for a summary of the distribution of clinician types (i.e., GP or nurse) across the question categories. Responses were not generated for the complete set of 5,609 categorised questions because a target of 5,000 questions was set, and clinicians were instructed to stop once that target had been reached (although there was some delay, hence the increased number of 5,422).

**mTable 1.** Percentage of questions answered by GPs and Nurses for each category.

| Category | % Answered by GPs | & Answered by Nurses | Total |
|---|---|---|---|
| Adolescent Sexual and Reproductive Health (ASRH) | 61.8% | 38.2% | 34 |
| Behaviour Change Communication (BCC) | 63.3% | 36.7% | 49 |
| Community-Based Provision Family Planning (CBP/FP) | 70.7% | 29.3% | 41 |
| Drug Management | 72.1% | 27.9% | 68 |
| Early Childhood Development (ECD) | 62.3% | 37.7% | 53 |
| Emergency Response | 57.7% | 42.3% | 26 |
| First Aid | 60.0% | 40.0% | 35 |
| Gender Based Violence (GBV) | 66.7% | 33.3% | 33 |
| HIV | 69.4% | 30.6% | 36 |
| Integrated Community Case Management (ICCM) | 74.5% | 25.5% | 51 |
| Malaria | 71.6% | 28.4% | 95 |
| Maternal & Newborn Health | 61.5% | 38.5% | 52 |
| Mental Health | 78.0% | 22.0% | 41 |
| Non-Communicable Diseases (NCDs) | 60.0% | 40.0% | 35 |
| Nutrition | 60.8% | 39.2% | 51 |
| Tuberculosis | 69.4% | 30.6% | 36 |
| Water, Sanitation, and Hygiene (WASH) | 57.5% | 42.5% | 40 |
| Other | 76.2% | 23.8% | 101 |

After providing their responses, clinicians were prompted to rate the quality of each vignette (on a 5-point Likert scale) for four criteria:

1. Relevance: how directly the question relates to the patient’s specific condition or the public health concern. Highly relevant questions enable clinicians to quickly grasp context and respond accurately.
2. Clarity: how easily the question is understood by healthcare providers.
3. Actionability: whether the question can clearly lead to practical steps in patient care.
4. Completeness: whether the question captures all essential information needed for clinicians to fully address the medical issue.

This rating step was included to capture additional insights regarding the quality and utility of the questions received from CHWs. We found that 90% of all vignettes received ratings of 3 or higher across all dimensions. This indicates that most questions submitted by CHWs were both relevant and adequately comprehensive in terms of providing the necessary information for clinicians to respond.

**mFigure 5.**
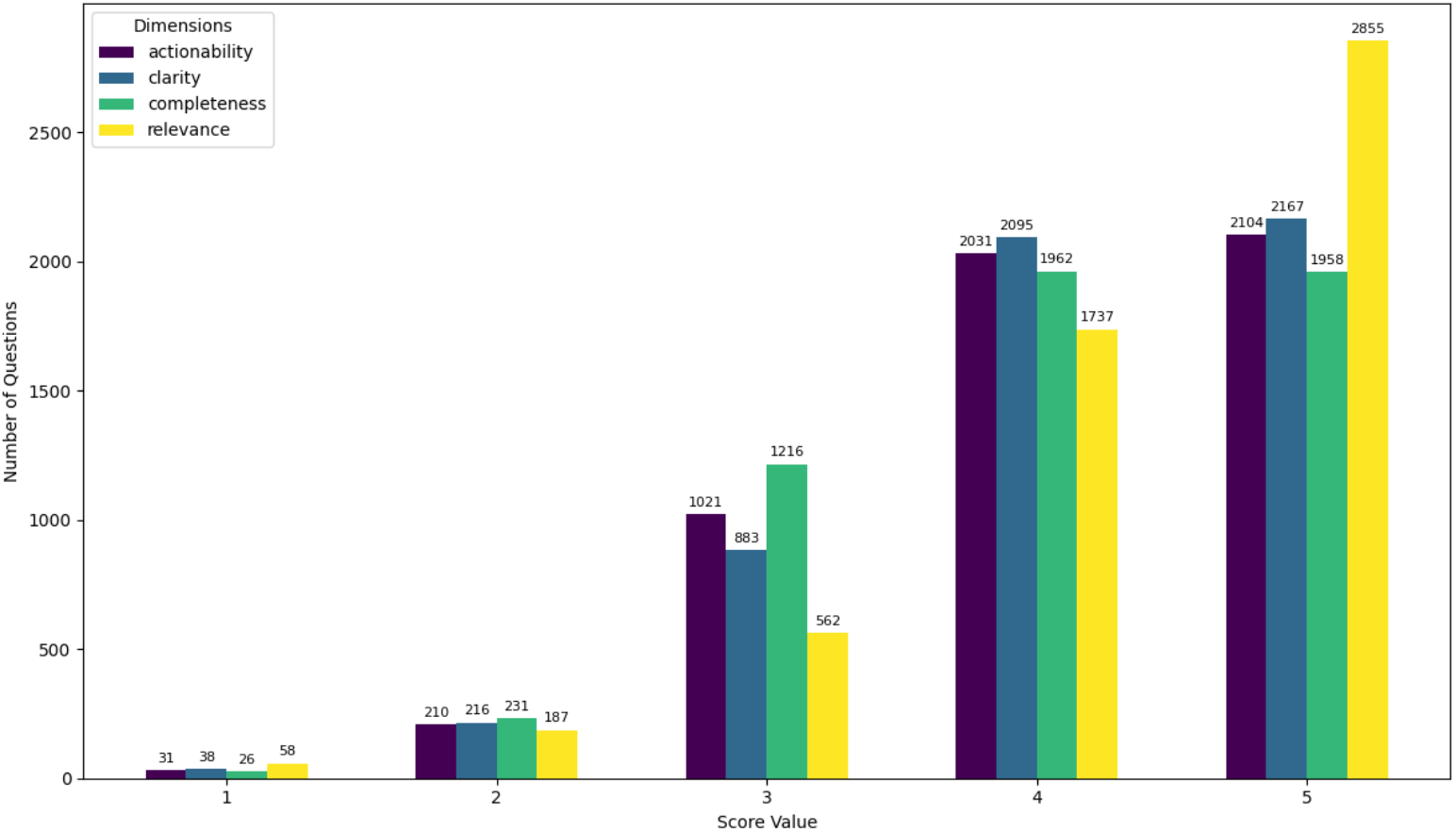
Histogram of the number of questions achieving each score for each of the four rating categories.

### LLM Response Generation

We generated responses to all 5422 vignettes in the final dataset from five LLMs: Gemini-2-Flash, GPT-4o, o3 mini, Deepseek R1, and Meditron-70B. For all but Meditron-70B, responses were generated using the relevant APIs by supplying the models with each vignette and the prompt shown in mBox 1. Since Meditron-70B is an open-source model, Digital Umuganda hosted the model in a private cloud instance (utilising two A100 GPUs within Google Cloud Platform), while maintaining the same prompting strategy.

**LLM Instruction Prompt**

**Role & Context**

You are a knowledgeable doctor,supporting Community Health Workers (CHWs) in Rwanda. You focus on providing accurate health advice, practical tips, and emotional support. You answer in the same language the question is asked (e.g., Kinyarwanda, English). You adapt to local conditions, respect cultural practices, and draw on global best practices. If uncertain, suggest ways to check information rather than giving incorrect details.

**Instructions**

Offer reliable medical guidance in a concise way.

Include local Rwandan health guidelines where possible.

If needed, give motivational or emotional support to CHWs.

Consider resource-limited settings and propose creative solutions.

Emphasize safety, fairness, confidentiality, and kindness.

All models, except Meditron, were prompted natively in English and Kinyarwanda, i.e., both the original Kinyarwanda transcription and the English transcription of each vignette were provided to solicit one response in each language from each model.

The cost of each response generated by each model was measured by applying relevant tokenisers (o200k_base for GPT-4o and o3 mini; LlamaTokenizerFast for Deepseek R1 and Meditron-70B; and Gemma’s SentencePiece-based tokeniser for Gemini-2) to tokenise all vignettes supplied as prompts and all responses received from each model. The input and output token counts were then combined with the per-token costs for each model to calculate the inference cost for each question-answer pair.

### Comparing Human and Model Performance

To compare the responses generated by local clinicians with those generated by the five LLMs included in this study, we recruited a panel of local experts to evaluate response quality, and then we analysed differences in how human clinicians and each of our models performed.

#### 1. Human Evaluation

Six clinicians were recruited to evaluate a set of 506 question-answer pairs. Clinicians were recruited by the Director of Clinical Services at Butaro District Hospital (Author ER), specifically targeting senior doctors with at least three years of clinical experience. They were paid 13,235 Rwandan Francs (~$ 9.17 USD) per question-answer pair evaluated. Question-answer pairs were sampled randomly to select 416 cases that would be evaluated in English (i.e., the vignette and the human/model-generated response would be presented in English), and 108 that would be evaluated in Kinyarwanda (i.e., the vignette and the human/model-generated response would be presented in Kinyarwanda). Since question-answer pairs were sampled at random, some of the 416 cases that would be evaluated in English were originally responded to by human clinicians in Kinyarwanda, with those responses later machine-translated into English (and vice versa for the 108 cases evaluated in Kinyarwanda) – this was accounted for in our analysis (see below).

Evaluating clinicians used an adaptation of the Med-PaLM-2 evaluation framework^1^ to evaluate each question-answer pair. The full evaluation framework used can be found in the ‘Supplementary Evaluation Framework Description’, but in brief, clinicians rated each response on 11 dimensions:

1. Alignment with Medical Consensus: *Does the response align with established medical guidelines, evidence-based practices, and expert consensus?*
2. Question Comprehension: *Does the response accurately understand and address the question asked?*
3. Knowledge Recall: *Is the information provided accurate, relevant, and reflective of an expert-level knowledge base?*
4. Logical Reasoning: *Is the response logically structured, with a clear and coherent rational progression of ideas?*
5. Inclusion of Irrelevant Content: *Does the response include unnecessary or unrelated information that could distract from the question at hand?*
6. Omission of Important Information: *Does the response omit any critical information that would compromise its quality, accuracy, or safety?*
7. Possible Extent of Harm: *If the user were to follow this response, how severe could the potential harm be (e*.*g*., *misdiagnosis, incorrect treatment, or unsafe advice)?*
8. Possible Likelihood of Harm: *How likely is it that the response could lead to harm if followed?*
9. Clear Communication: *Is the response presented in a clear, professional, and understandable manner? Is the structure and tone appropriate for the intended audience?*
10. Understanding of Local Context: Does the response take into account regional, cultural, and resource-specific factors relevant to the local setting in Rwanda?
11. Potential for Demographic Bias: *To what extent does the response avoid bias based on demographic factors such as age, gender, race, ethnicity, or socioeconomic status?*

The evaluation itself was conducted by dividing the clinicians into two groups of three clinicians each, with one clinician in each group designated as a supervisor and the other two as evaluators. Each group assessed 262 question-answer pairs (i.e., half of the full sample). Initially, evaluators would independently evaluate six responses (i.e., the five models and one human response) generated for the same question. They would then convene to discuss their evaluations and resolve any disagreements in their scoring (disagreement defined as a difference of >1 on the 5-point Likert scale for each dimension). This process yielded two sets of independent scores for each response generated for each vignette, with each pair of per-dimension scores being within one Likert-point of one another. Disagreement could not be resolved for 15 English question-answer pairs and 3 Kinyarwanda question-answer pairs; these cases were removed from the dataset and all subsequent analyses.

#### 2. Statistical Analysis

The primary question we sought to answer was whether and how the quality of responses depended on the source, specifically, how humans and the five models compared, as evaluated by expert human clinicians. We also sought to understand whether the profession of the responding clinicians (i.e., whether they were senior nurses or junior GPs), the language used to respond (i.e., whether nurses/GPs responded in English or Kinyarwanda), or the language used to evaluate (i.e., whether evaluating clinicians evaluated English or Kinyarwanda question-answer pairs) had any effect on performance.

To answer these questions, we conducted an Aligned Rank Transform (ART) ANOVA^14^, which is a non-parametric factorial procedure that “aligns” the data by subtracting estimated effects for each term, then ranks the aligned values so that a standard ANOVA performed on those ranks yields valid tests of main effects and interactions without assuming normality or interval scaling. Because alignment isolates each effect before ranking, the method maintains Type I error control and statistical power comparable to that of parametric ANOVA, even with small samples or skewed, ordinal outcomes (such as the Likert-scale rating used in our evaluation framework).

We tested a fully saturated statistical model, which included main effects for evaluation dimension (i.e., which of the 11 evaluation dimensions an individual score pertained to), responder (i.e., which of junior GP, senior nurse, GPT-4o, o3-mini-high, Gemini-2-Flash, Meditron-70B, or Deepseek-R1 generated the response under evaluation), and evaluation language (i.e., the language of the question-answer pair presented to evaluating clinicians), along with all interaction terms.

We then conducted pairwise comparisons between the levels of all significant main effects and interactions on the aligned-ranked marginal means with Tukey’s honest significance difference (HSD) test. Because the ART procedure isolates each factorial contrast before ranking, the aligned ranks meet the independence and equal-variance requirements of HSD, allowing us to control the family-wise error rate. This approach yields adjusted p-values for all pairwise contrasts within each significant main effect or interaction, providing a rigorous yet interpretable basis for reporting differences we found among evaluation dimensions, responders, and languages.

Finally, we analyzed differences in the cost of generating responses in English versus Kinyarwanda for the three models that could do so (excluding Meditron-70B and Deepseek, as they were unable to operate natively in Kinyarwanda). For all API-accessed models, the cost of each response was computed by tokenizing the input and output text with the appropriate tokenizers and then applying the input/output token costs to the resulting number of tokens. Costs in cents per million tokens, for input and output respectively, were: GPT4o (2.5, 10), Gemini-2-Flash (0.5, 2), o3 Mini-High (1.1, 4.4), and DeepSeek R1 (0.55, 2.19). For humans, the cost of each response was the amount paid to the junior GP or senior nurse who generated it. Costs were analyzed using a standard two-way ANOVA, which included main effects for model and language, as well as the interaction between them, all of which were significant. In brief, we hypothesized that generating responses in Kinyarwanda would be more expensive due to larger token counts for the same semantic content (when compared with English translations of the same content).

The complete set of analytical code is archived in a fully open repository, available at: https://github.com/PATH-AI-Initiative/RwandaBenchmarking

## Ethics Approval

The study was deemed exempt from review by the Rwanda National Ethics Committee. PATH’s research determination committee also reviewed the scope and confirmed it was not human subjects research subject to IRB approval.

## Data Availability Statement

The subset of 524 questions, answers, and individual evaluation results that comprise this benchmarking study is available upon reasonable request and will otherwise be made available upon publication of this work in a peer-reviewed journal. The full dataset has been donated to the Rwanda Biomedical Centre (RBC), the parastatal delivery arm of the Rwandan Ministry of Health, and is hosted in a secure data environment. It will be made available to researchers on request and based on an assessment of ‘fair value exchange’ by stakeholders, to ensure that the indigenous population that generated the information benefits from its exploitation. This arrangement was specifically designed to ensure adherence to the CARE principles. The Centre for the Fourth Industrial Revolution, as the innovation lab for the Rwandan Government, serves as the primary point of contact for researchers seeking to access this data. Prospective users should contact ‘’ to request access.

## Patient and Public Involvement Statement

Patients were not directly involved in this study.

## Author Contributions Statement

BAM conceptualized the study, and secured funding for it. CN and MEF managed the project. SR, GW, CN, MEF, VM, XL, AD, and BAM developed the methodology. EH, SR, KK, FN, SU, ER and other members of the broader Digital Umuganda team (acknowledged) developed the tools, recruited relevant participants, for the study and collected the data. SR, GW, and members of the broader Digital Umuganda team (acknowledged) performed the data analysis. SR, MEF, GW, and BAM drafted the original manuscript. All authors contributed to review, editing and approval of the final manuscript.

## Competing Interests Statement

The authors declare no competing interests.

## Funding Statement

This research was supported by the Gates Foundation (INV-068056). The funders had no role in the study design, data collection and analysis, the decision to publish, or the preparation of the manuscript.

## Acknowledgments

We thank the Ministry of Health and the Ministry of ICT for their support, as well as the numerous community health workers (CHWs) and clinicians who participated in the study. Additionally, we’d like to acknowledge the efforts of the broader Digital Umuganda team (Boris Ishimwe Mugisha, Michel Mivumbi Patrick, Saad Byiringiro, Celestin Niyindagiriye, Cedric Mugisha, Ali Nengo, Emmanuel Igirimbabazi, Gilbert Nzabonimpa), as well as the C4IR leadership (Crystal Rugege & Alain Ndayishimiye) for their contributions in operationalizing and undertaking this study.

## Notes

### Competing Interest Statement

The authors have declared no competing interest.

### Author Declarations

IRB of Rwanda National Ethics Committee waived ethical approval for this work.

## References

1. Singhal K, et al. Nature Medicine. 2025;, Jan 8:1–8.

2. Goh E, et al. Nature Medicine. 2025;, Feb 5:1–6.

3. Idriss-Wheeler D, et al. PLOS global public health. 2024;, Jan 18;4(1):e0002799.

4. Olatunji T, et al. Available at: https://github.com/intron-innovation/AfriMed-QA. Accessed on 24 May 2025;.

5. Olatunji T, et al. arXiv preprint arXiv:2411.15640. 2024;, Nov 23.

6. Elamin M, Chanie Y, Ewuzie P, Rutunda S.. In 4th Workshop on African Natural Language Processing 2023; Apr.

7. Alhanai T, et al. In Proceedings of the AAAI Conference on Artificial Intelligence 2025;, Apr 11.

8. Ahia O, et al. arXiv preprint arXiv:2305.13707. 2023;, May 23.

9. Johri S, et al. Nature Medicine. 2025;, Jan 2:1–0.

10. Menon V, et al. Available at: 10.5281/zenodo.15493572

11. Mateen BA, Nature Medicine. 2025;, [In Press].

12. Joint Commission, 2008;. available at https://www.jointcommission.org/at_home_with_the_joint_commission/sbar_%E2%80%93_a_powerful_tool_to_help_improve_communication/. Accessed 24 May 2025;.

13. Digital Umuganda. Hugging Face. 2024;. Available at: https://huggingface.co/DigitalUmuganda/Quantized_Mbaza_MT_v1

14. Wobbrock, J.O., Findlater, L., Gergle, D. and Higgins, J.J., In Proceedings of the SIGCHI conference on human factors in computing systems. 2011;.

